# Primary care clinicians working in or near hospital emergency departments in the UK: A mixed methods systematic review

**DOI:** 10.1101/2024.08.19.24312212

**Authors:** Stephanie Howard Wilsher, Julii Brainard, Sarah Hanson, Duncan Peacock, Paul Everden

## Abstract

**Objectives:** To synthesise evidence about primary care clinicians treating patients in or adjacent to hospital emergency departments in the UK.

**Study design:** Mixed methods narrative systematic review.

**Methods:** Eligible studies were in English and described primary care services (general practitioners, GPs, or nurse practitioners) that treat patients within or adjacent to hospital Emergency Departments (ED). Searches were conducted on Medline, EMBASE, Cochrane Library and CINAHL databases. The search included extraction from an international review updated from 2020 to October 2022, and grey literature from inception to October 2022. The methods were informed by consultation with members of the public.

**Results:** From 4189 studies screened, 20 met inclusion criteria. Four studies assessed typology and streaming of services. Seven studies reported patient and public involvement. Ten studies reported differences in clinical outcomes between primary care and emergency services, but not definitive benefit for either. Likewise, results were equivocal for economic evaluations. Diverse delivery formats complicate evaluation and may explain why clinicians had mixed opinions about the utility of such services. Patients were generally satisfied with the service they received, in either primary care or emergency services.

**Conclusion:** Diversity of implementation complicate conclusions that can be drawn. Existing evaluations provide little evidence that primary care services in or near emergency departments offers any system advantages for clinical outcomes, or cost savings. Process evaluation in future evaluations is essential to understand what aspects of primary care at emergency departments are likely to improve system and patient care.

**Implications for the profession and/or patient care:** The systematic review assessed outcomes arising due to primary care clinicians providing treatment in or near Emergency Departments (ED) in the UK. There were no consistent benefits for having primary care clinicians in or near EDs. This research highlights the need for healthcare leaders and policy makers to provide more clarity in developing primary care services in or near EDs. It will have impact for leaders in healthcare to consider whether primary care clinicians in or near EDs are providing ‘best value’ healthcare, or whether other models could provide patients appropriate NHS resources according to their health need.

**Patient and public involvement:** Three focus groups (with 13 public advisors) were conducted to understand patients’ priorities and perspectives for attending EDs with relatively minor health conditions. This helped to guide study design, data extraction and analysis of this review.

## Introduction

Unscheduled health care seeking at NHS emergency departments (EDs) has been growing at a pace that exceeds population growth (Baker, 2017). Many interventions to improve patient flow and to meet service performance targets, have been tested and continue to be trialled in the NHS. Before the pandemic, estimates were that 15-44% of Emergency Department (ED) attenders in Britain present with conditions that can be addressed by primary care clinicians (Ismail et al., 2013; O’Keeffe et al., 2018). In response, embedding primary care clinicians into care pathways available to ED attenders has become widespread practice in the NHS. Our systematic review seeks to assess the efficacy of primary care in or near ED’s in the UK.

A recent systematic review (searches through January 2020) scoped interventions where patients were triaged and/or treated by primary care professionals to reduce overcrowding in EDs globally (Jeyaraman et al., 2021). Many outcomes, time to provider initial assessment, time to triage, proportion of patients leaving without being seen, number of repeat ED visits, and patient satisfaction, favoured the primary care initiatives. However, the UK has some distinctive urgent care system features. The NHS provides free care at point of delivery.

Whether payment is required (or not) at care delivery point is known to distort health care seeking choices in other settings (Pekerti et al., 2017; Reich and Shibuya, 2015). NHS EDs are expected to discharge, admit or refer 95% of all service users within four hours of presentation (Mortimore and Cooper, 2007). There are potential financial penalties levied on NHS providers for failing to meet this target, thus the 4-hour key performance indicator (KPI) itself is thought to distort patient experiences and care pathways, potentially to patient detriment (Black, 2022).

We were interested in trying to summarise evidence about potential service benefits where the additional care pathway was likely to be truly primary care as opposed to provision of additional professionals to treat high numbers of arrivals some of whom are low acuity. We took this perspective because of the possible differences in diagnostic strategy and risk management that may distinguish primary care (Heneghan et al., 2009) from ED (Medford-Davis et al., 2018) clinicians. Our data extraction therefore was not simply about service activity and system impacts, but also considered aspects of whether appropriate care (proportionate and not escalating, rather than over-investigation or over-treatment) was provided.

This systematic review was prompted by the merits of considering the unique aspects of urgent health care provision in the UK as well as publication (since 2020) of several UK-focused original research studies about primary care health care pathways at EDs. We adopted a holistic approach to cover all aspects of primary care in or near EDs in the UK; thus, we expected high heterogeneity in the relevant studies. To accommodate the variations in study subject matter, a mixed method, narrative review study design was the most appropriate (Mays et al., 2005).

## Methods

The systematic review was conducted in line with Cochrane Review guidelines (Higgins, 2022). A protocol was developed to guide the systematic review. We benefited from a previous scoping review (Jeyaraman et al., 2021) that had overarching inclusion criteria, in that it included initiatives that involved primary care staff in streaming or treatment of arrivals at EDs. We therefore screened all included studies in Jeyaraman et al. (2021) for potential inclusion in our review, as well as adapted their search strategy to find additional eligible studies published after their last search date (January 2020) and by our search date, 10 October 2022.

### Patient and public involvement

Our synthesis was informed by parallel public and patient involvement (PPI) undertaken in three focus group discussions with community members. Among other topics, our groups discussed patient priorities and perspectives when deciding whether to attend emergency departments with relatively minor health conditions. The PPI advisors described determinants of patient satisfaction which helped to inform our choices of what information to extract and summarise in this review.

### Ethical Approval

Ethical approval was not required for PPI as the community members were advisors to inform our research. Likewise ethical approval was not required for the systematic review as it included secondary data.

### Clinical carers

It was unexpectedly challenging to define who is a primary health care professional and who is not. The distinction mattered because our focus was on exactly who delivered treatment or care, rather than if the clinician operated only in a triage role. We wanted to focus on possible merits of service models which involved treatment being delivered by ‘generalists’ rather than specialists. We ultimately defined primary carers as individuals who were described as general practitioners or as nurse practitioners (NP) or advanced NP, when *not* acting in the role of emergency nurse practitioner (ENP). This distinction (about functional role of ENPs) was made because the role of ENP only emerged in the mid-1990s. In early articles, ENP was often described as a slightly enhanced role for nurse practitioners who were being asked to work according to ED protocols and diagnostic options. ENP since then has become its own role with unique qualifications. To exclude from our review NPs and only include GP-administered care was not desirable because many primary-care-at-ED initiatives in the UK feature a mixed staff of GPs and NPs. Under advice of co-authors PE (experienced GP) and DP (ED consultant), it was agreed that nurse practitioners placed in an ENP role would deliver care more in line with protocols expected of ENPs than ANPs working in a purely primary care setting. Therefore, any ANPs put in role of ENPs we treated as emergency care specialists, not primary care clinicians. We were advised, however, that the very different (and long) training that British GPs experienced made them much more likely to ‘think like a generalist’ and not like an ED specialist. Therefore, GPs were classified as primary care clinicians regardless of how they were integrated into urgent care pathways.

### Eligibility criteria

A participant, intervention, comparator, outcomes, study design (PICOS) table was designed to identify appropriate studies. Inclusion criteria required studies to be conducted in the UK where primary care clinicians (GPs, NPs or ANPs) provided treatment to patients who presented in person to EDs, Table 1.

**Table 1:** PICOS table developed for the systematic review on primary care in Emergency Departments.

|  |  |
| --- | --- |
| **Population** | **Any patient population (children and adults of any age) visiting ED where GDAE services are offered** |
| **Exposure/Intervention** | Treatment provided by primary healthcare professionals (GP, NP or ANP) acting in a primary care treatment role (not merely providing triage, not providing treatment at same point as triage, and not substituting as ENPs or other emergency/urgent care providers). |
| **Comparator (if data are available)** | Other treatment pathways (“usual care”) for ED attenders |
| **Outcomes** | All ED clinical outcomes that reflect the impact of primary care clinicians providing treatment (e.g., proportion of patients that leave ED without being seen (LWBS), ED length of stay (LOS), mean/median patient wait times in ED, percentage of patients with reduced wait times in ED, prescription, referral rates, and volume of patients visiting ED), re/admission to ED). Patient satisfaction and staff views of the service. Cost evaluations |
| **Study design** | All study designs will be included which record original data about patient or staff experiences because of patients who present for Face-to-Face non-urgent care and are provided it almost immediately on site. |
| **Publication** | Published studies and grey literature |
| **Date of studies** | Database inception to October 2022 |
| **Publication** | Published studies and grey literature |
| **Language** | English only |

### Data sources

Updated searches were conducted on Medline (Ovid), EMBASE, Cochrane Library (Wiley) and CINAHL (EBSCO) databases from January 2020 to October 2022, as well as screening all 268 studies included in the Jeyaraman et al. (2021) review.

### Search strategy

A published search strategy (Jeyaraman et al., 2021) with last search date January 2020 was adapted as this recent publication had inclusion criteria that encompassed our own inclusion criteria. Our own search (Table 2) was run to include additional searches for studies published January 2020 to October 2022. Reference lists of included studies were also checked for any studies that were not found by the searches. Grey literature was sought from inception to December 2022, using BMJ Open Quality and a Google custom search of the following websites: Agency for Healthcare Research and Quality, NHS Improvement, and International Society for Quality in Health Care.

**Table 2:** Inclusion/exclusion criteria for study selection for the systematic review on primary care in Emergency. Departments

| Inclusion criteria | Exclusion criteria |
| --- | --- |
| Face-to-face visits | Telemedicine |
| Patients receive healthcare in unplanned care settings where emergency/urgent treatment is an immediate alternative/usual care pathway | Patients for whom no other pathway exists at the setting |
| Patients receiving treatment from primary care staff (NPs, GPs, ANPs) in an unplanned, within/adjacent to ED clinical setting | Patients who receive non-urgent treatment from emergency care professionals |
| Patients who receive care after triage from primary care clinical staff | Patients who only receive care at same time point as triage |
| GPs, NPs, ANPs providing care | ANPs asked to 'act up' as Emergency nurse practitioners in ED, or in a role that would be recognised in 2022 as that of ENP, even if the role was described as ANP in the past (typically in 1990-2005 documents). Other types of health care professionals such as mental health nurses, physiotherapists, pharmacists, etc. are ineligible. |
| UK studies | Studies from other countries |

### Study selection

Studies were screened against the inclusion and exclusion criteria, Table 3. Two reviewers (SHW and JB) independently screened all abstracts identified through the search strategy and then conducted independent full text screening on the selected studies. Any disagreements were moderated by other research team members (SH, PE, or DP).

**Table 3:**
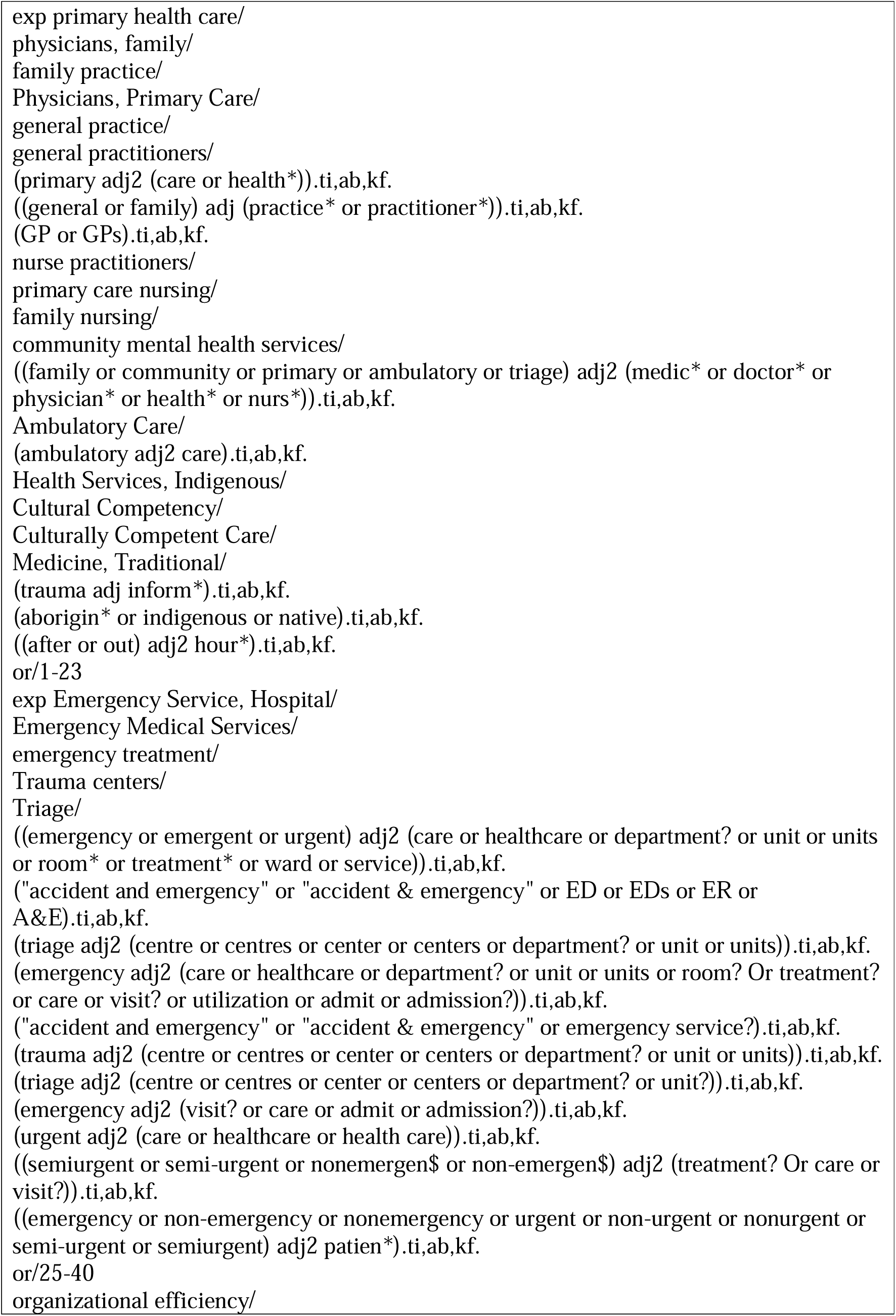
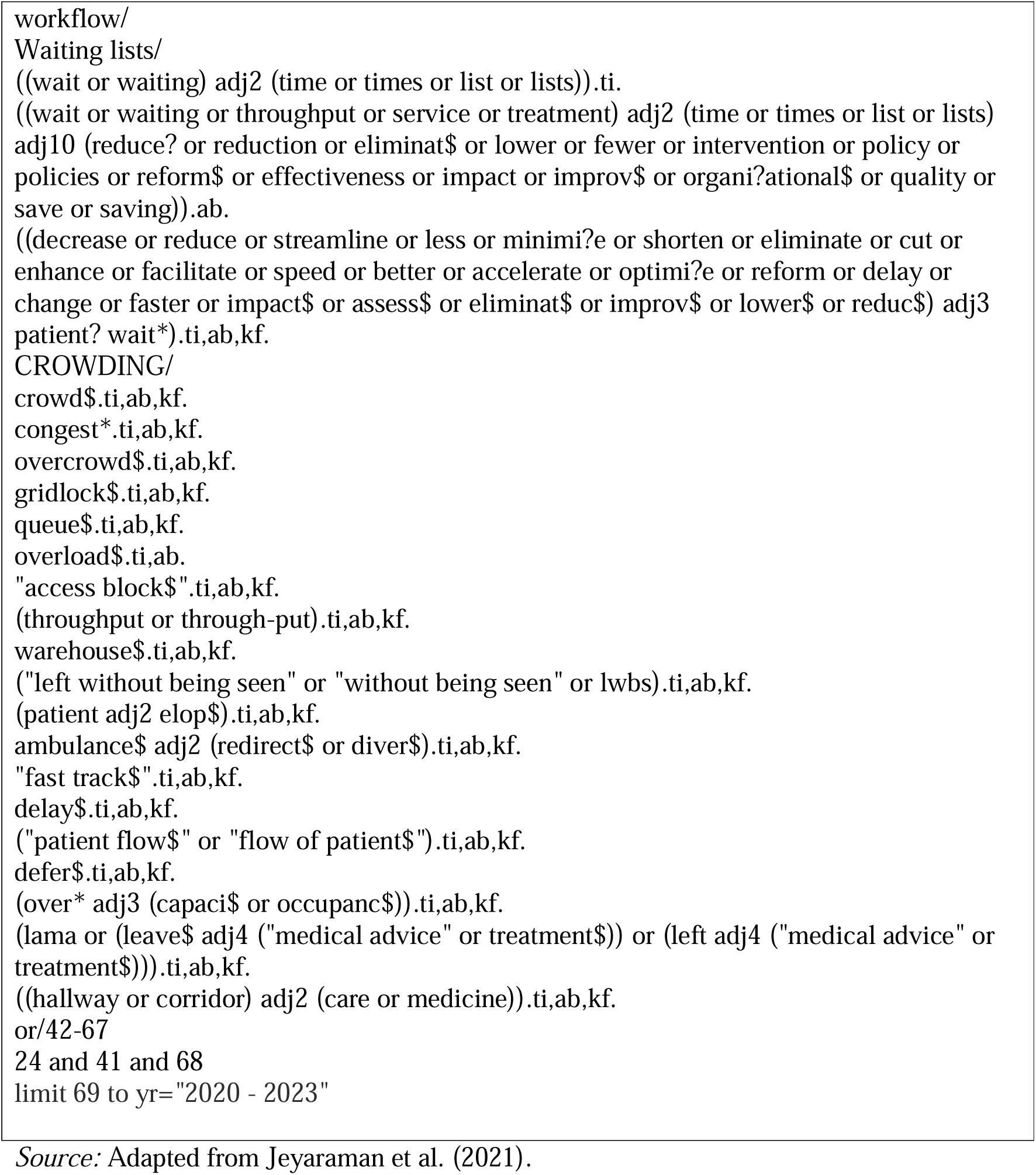
Search strategy on Medline (Ovid) for the systematic review on primary care at ED.

### Data extraction

Data were extracted by one reviewer (SHW) and checked by JB or SH. Data extracted included details of the service; study design; PPI activity (if any), taxonomies described (if any); types of streaming (if any); clinical outcomes (wait time; investigations, attendance counts; prescriptions; statistics about patients leaving without being seen; reattendance), adverse outcomes; patient experiences and views; staff perceptions; and economic outcomes.

### Synthesis methods

A mixed methods narrative systematic review was used with descriptive statistics. The findings were synthesised to provide a comprehensive overview of research on primary care treatment in or near EDs in the UK.

### Quality assessment

Quality assessment of included studies was determined with the Mixed Method Appraisal Tool (MMAT; Hong et al., 2018). MMAT was selected to cover the range of methodologies included in the review and has five questions pertinent to potential bias for each methodology. Studies were categorised as qualitative, quantitative, or mixed methods by the reviewers based on the evidence presented in the paper, rather than on the authors descriptions. Two reviewers (JB and SHW) independently conducted the quality assessment. Disagreements were discussed including with a third reviewer (SH) to reach consensus.

## Results

### Study selection

From 4189 studies identified in the searches, 20 met the inclusion criteria, study selection procedure is shown as Figure 1.

### Study characteristics

Study characteristics are shown in Table 4. All studies were about primary care clinicians treating patients in or near EDs. Five studies used mixed methods (Aldus et al., 2022; Coleman et al., 2001; Jones, 2011; Scantlebury et al., 2022; Brant et al., 2021), ten studies were quantitative (Dale et al., 1995; Dale et al., 1996; Gaughan et al., 2022; James et al., 2019; Leigh et al., 2021; McCarron et al., 2019; Salisbury et al., 2007; Uthman et al., 2018; Ward et al., 1996; Chalder et al., 2007), and five were qualitative (Ablard et al., 2017; Anderson et al., 2021; Edwards et al., 2020; Edwards et al., 2022; Edwards et al., 2021).

**Table 4:**
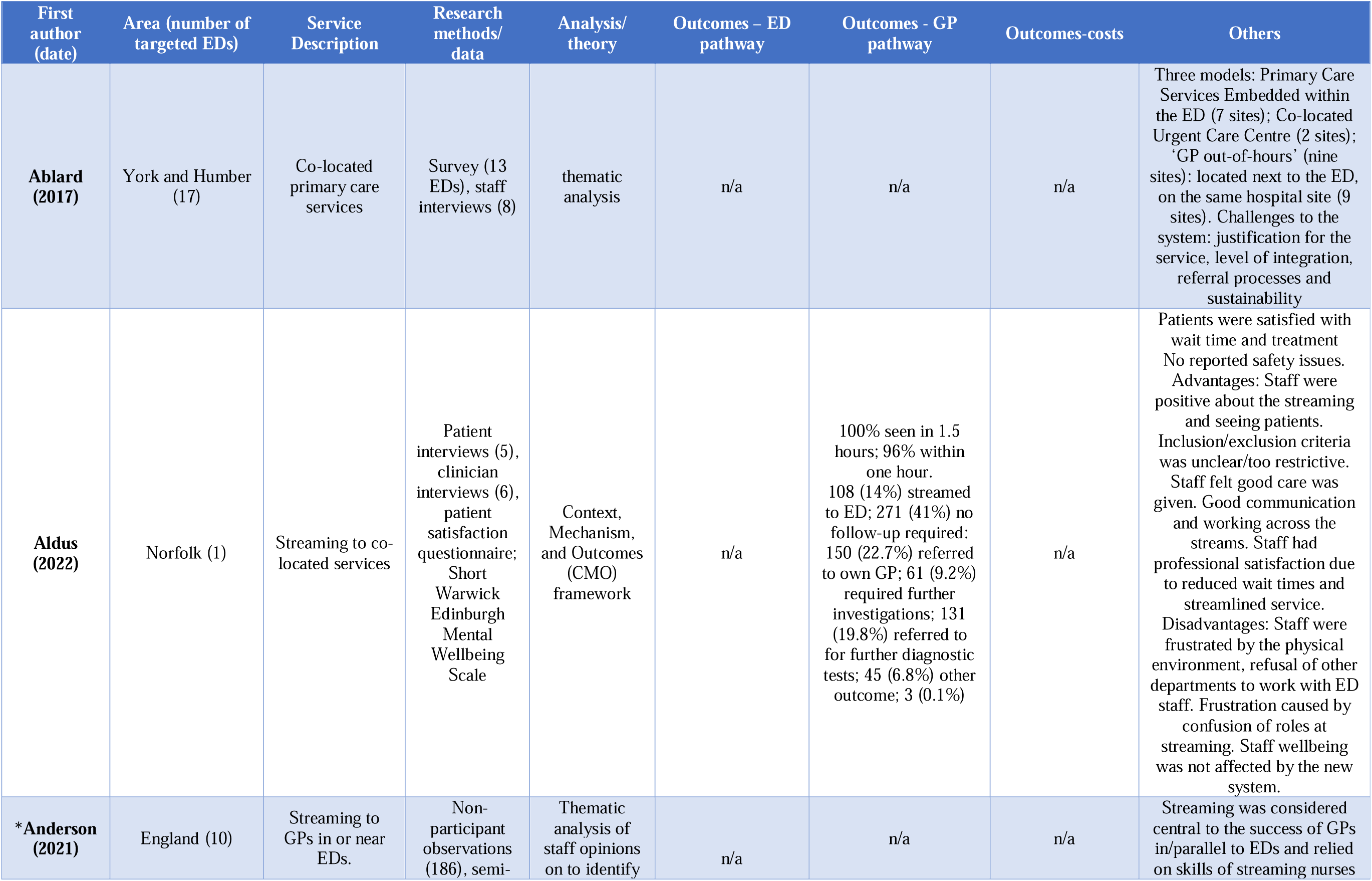
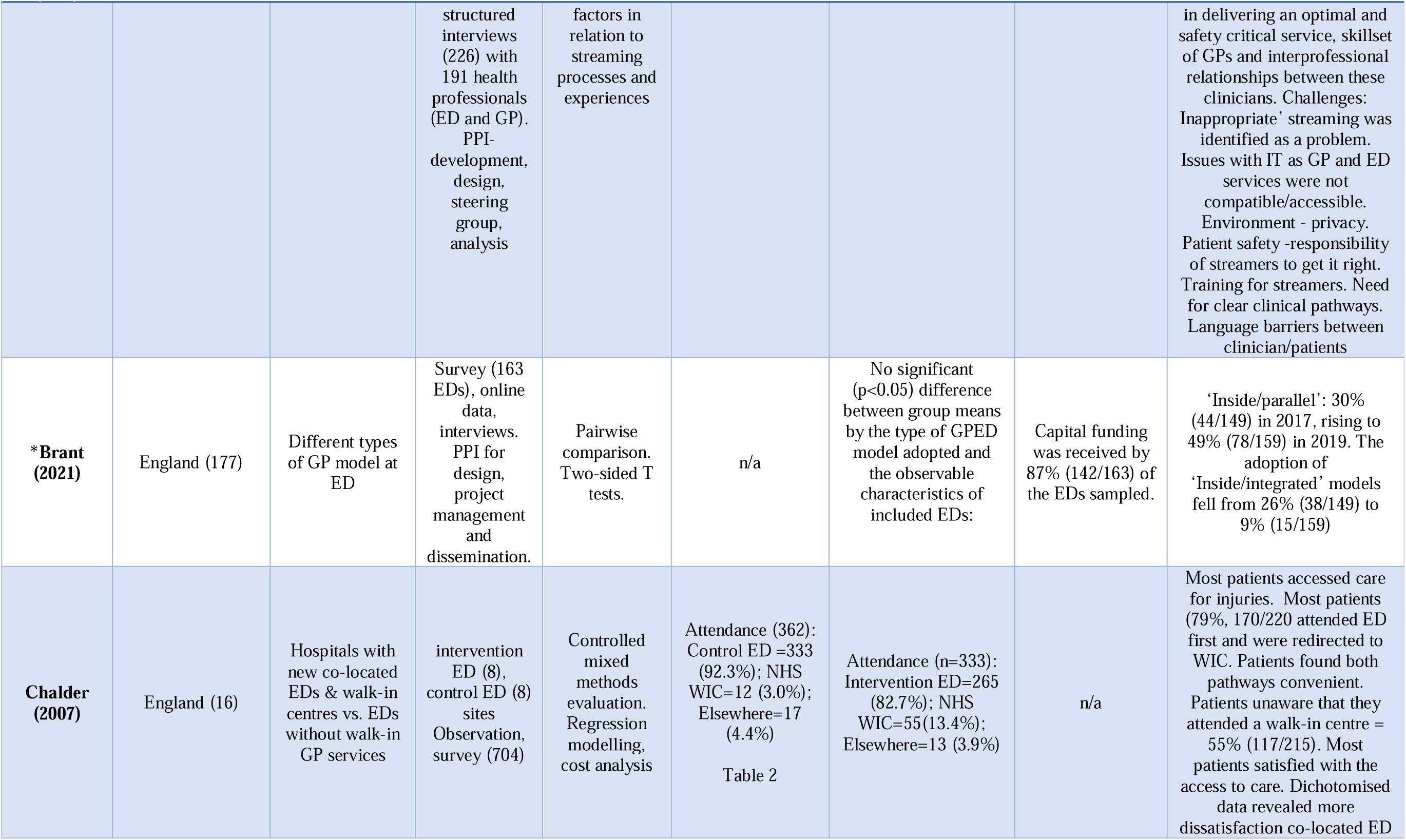
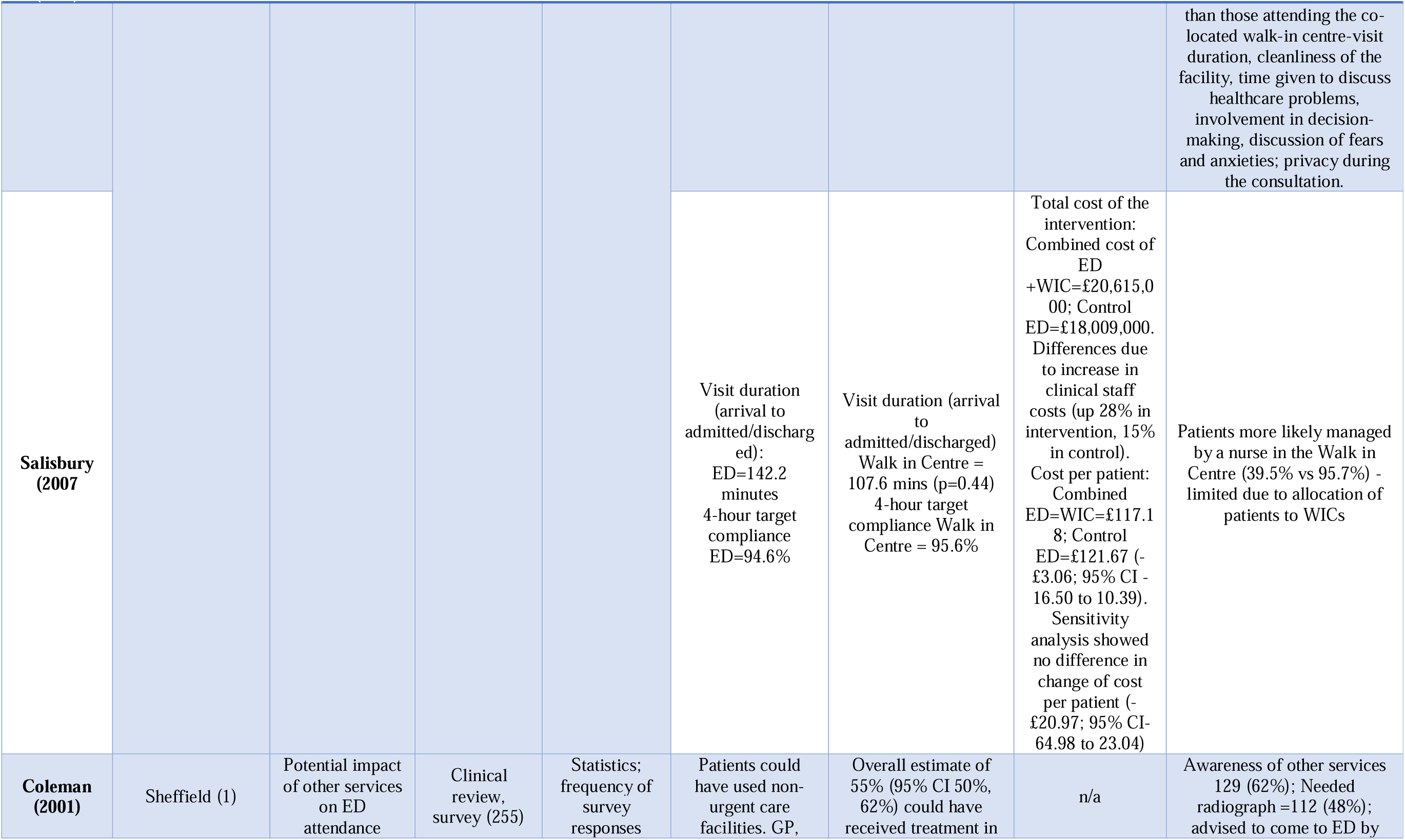
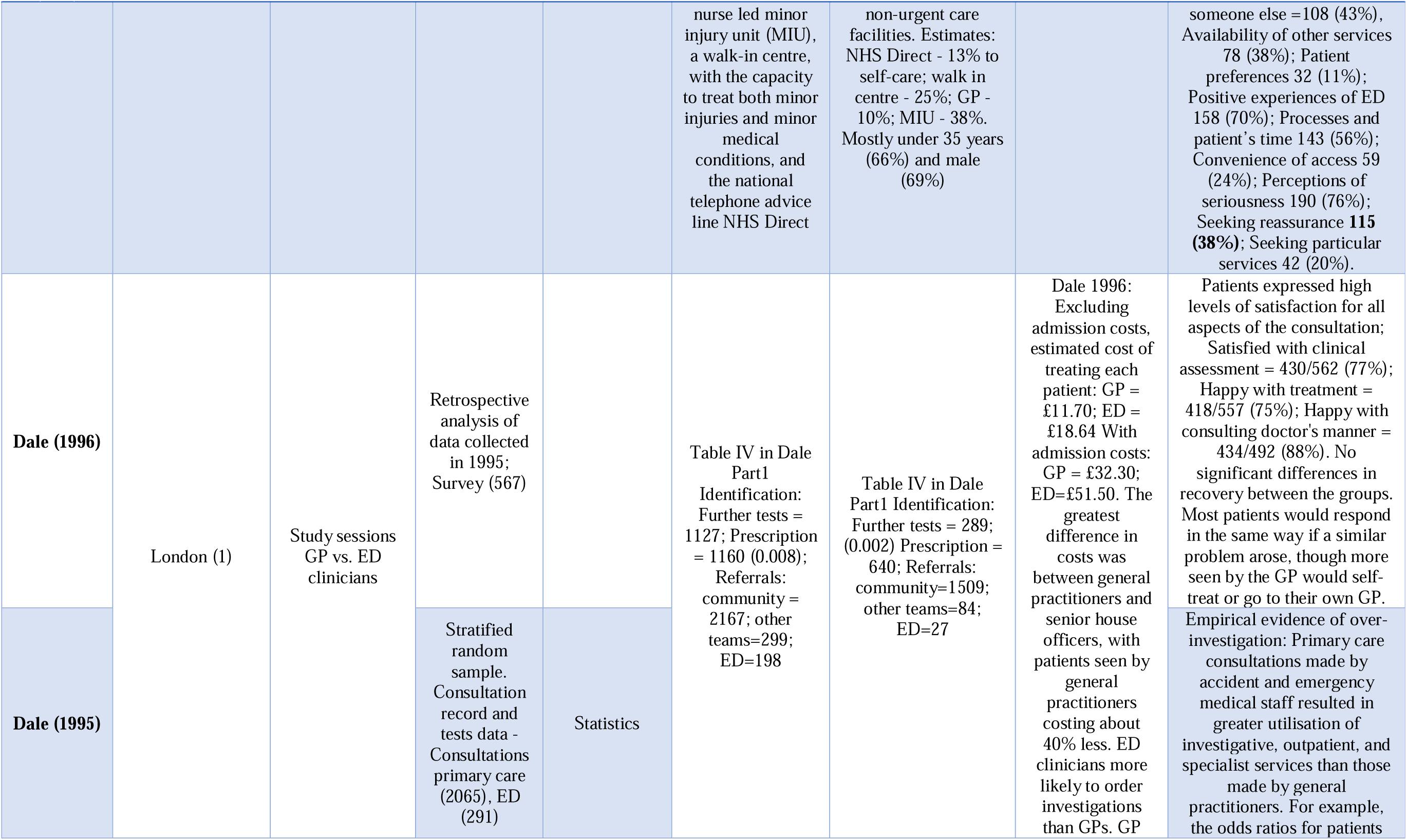
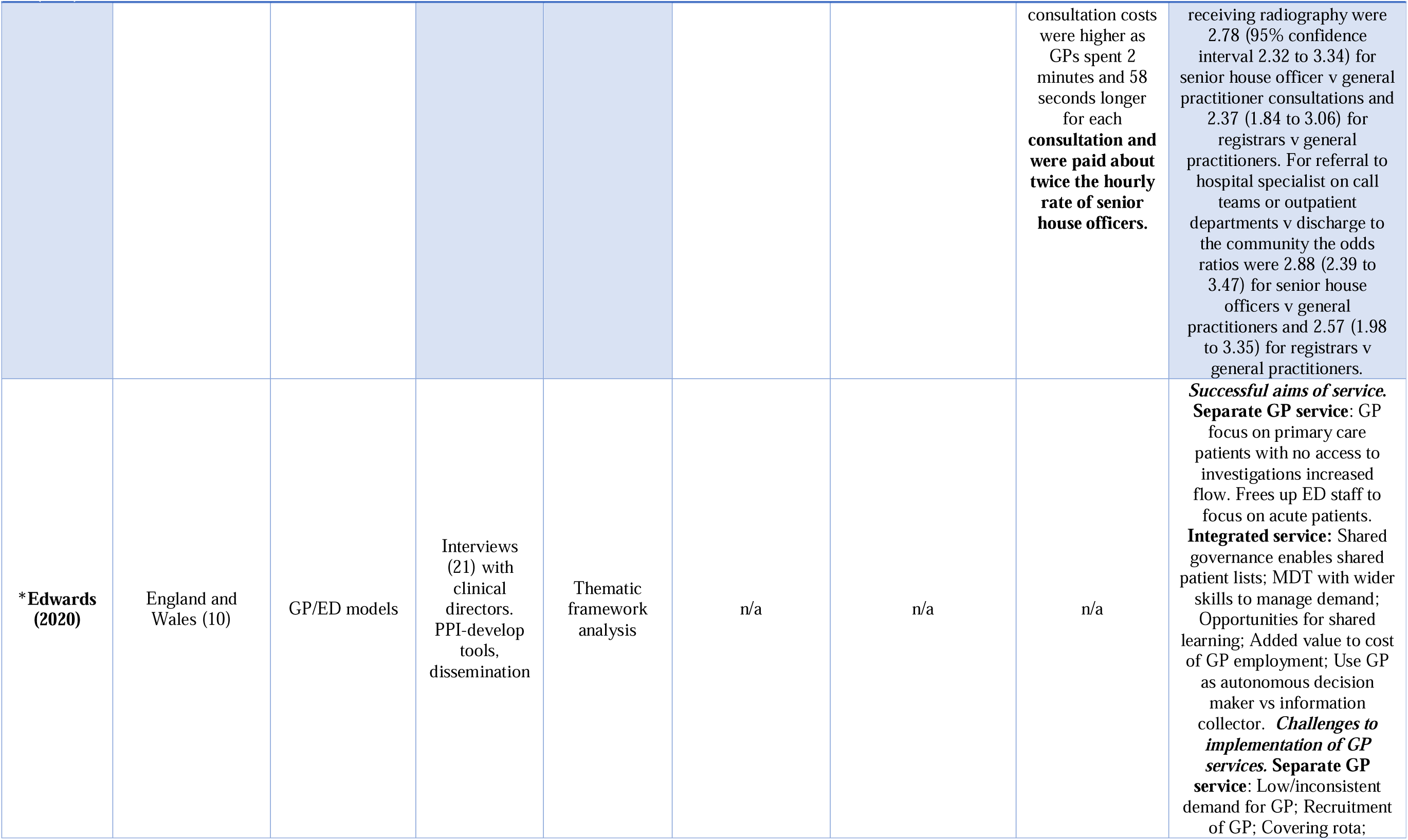
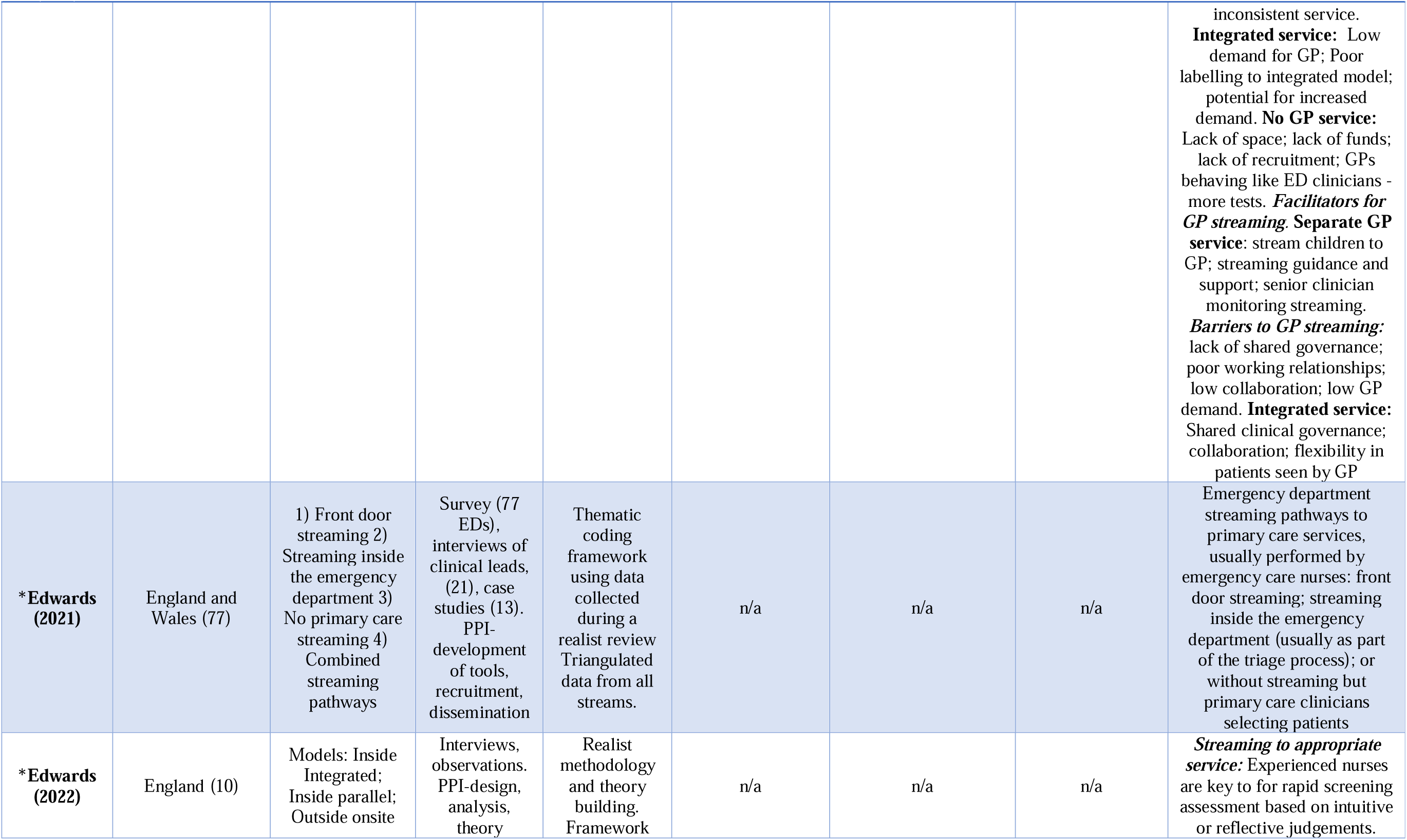
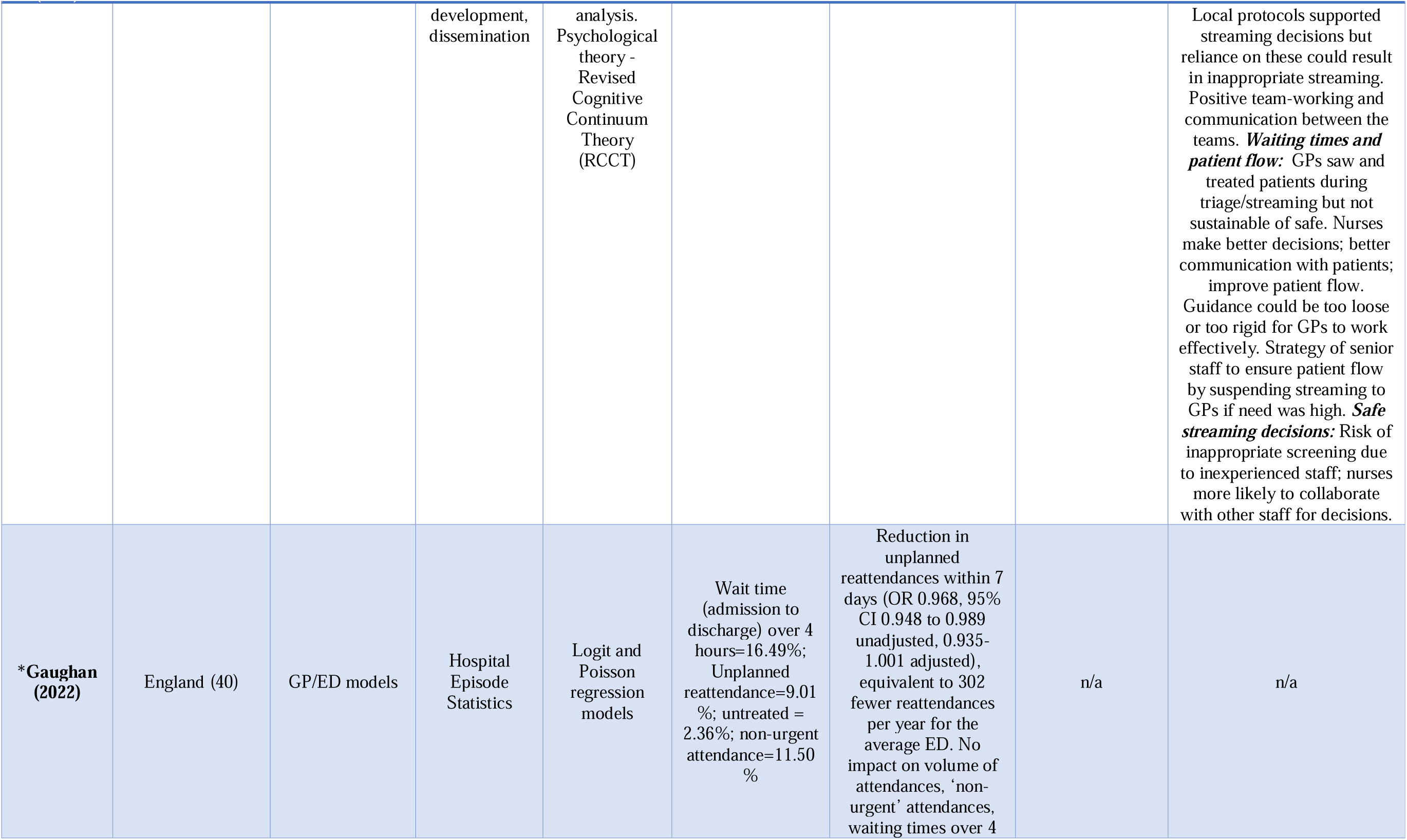
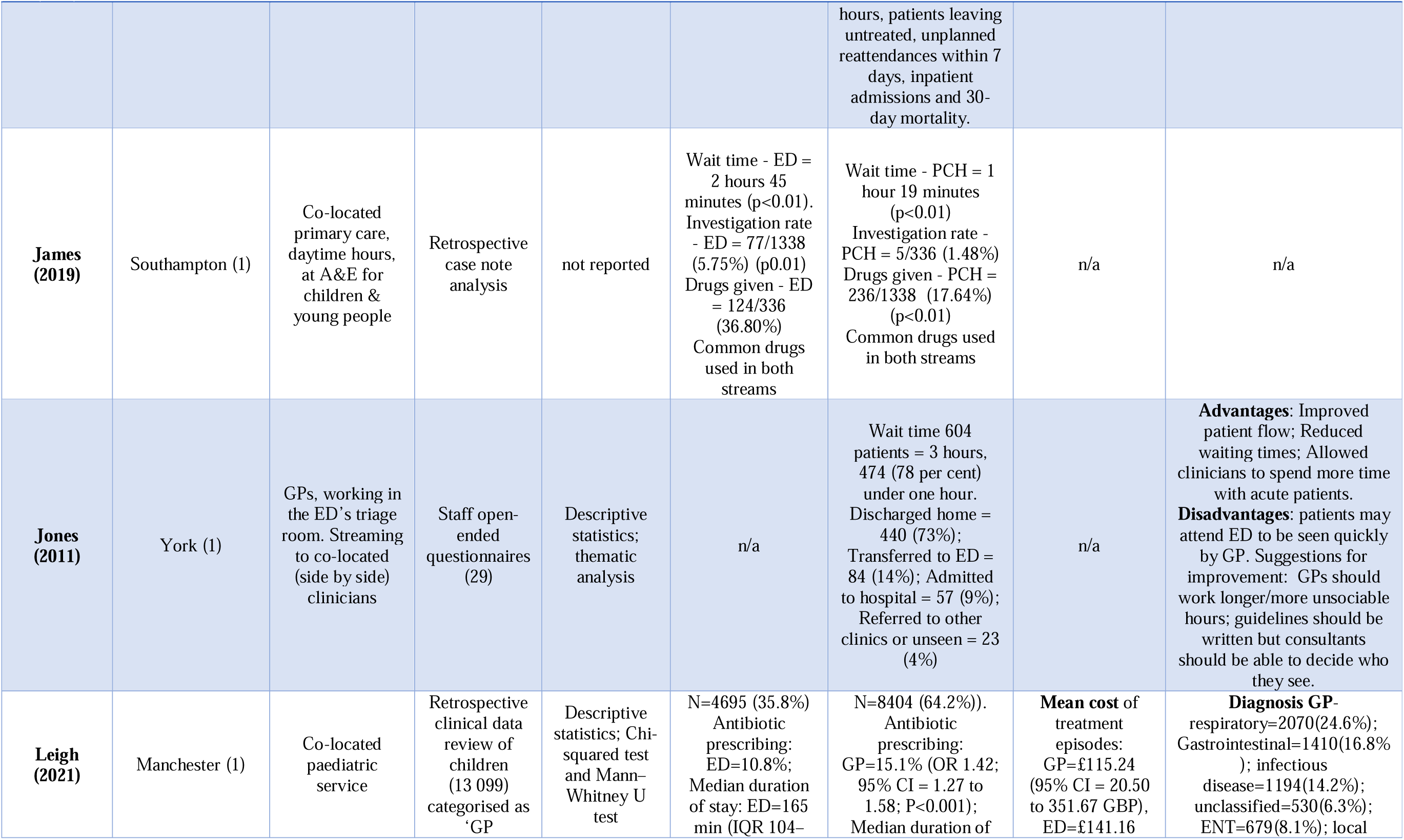
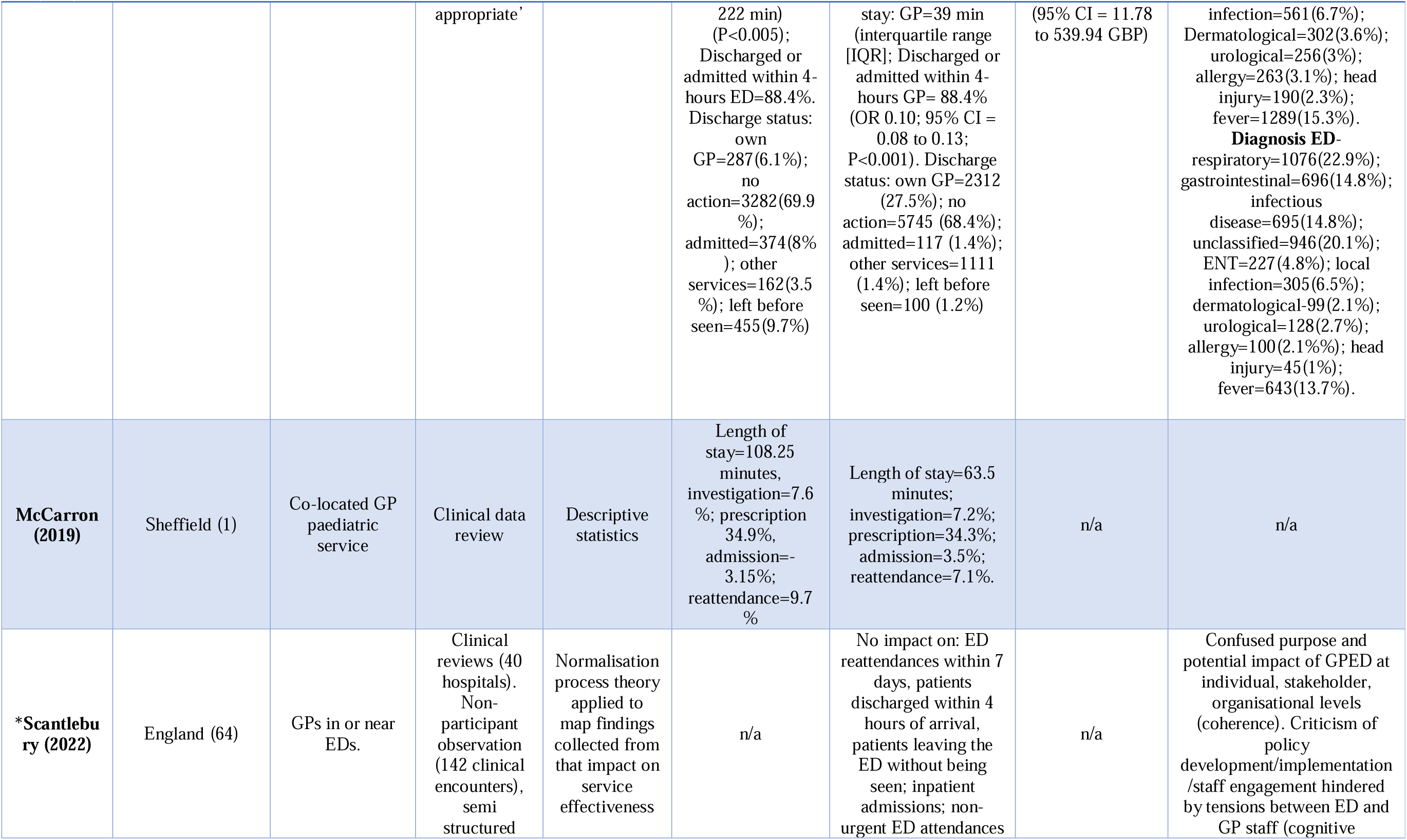
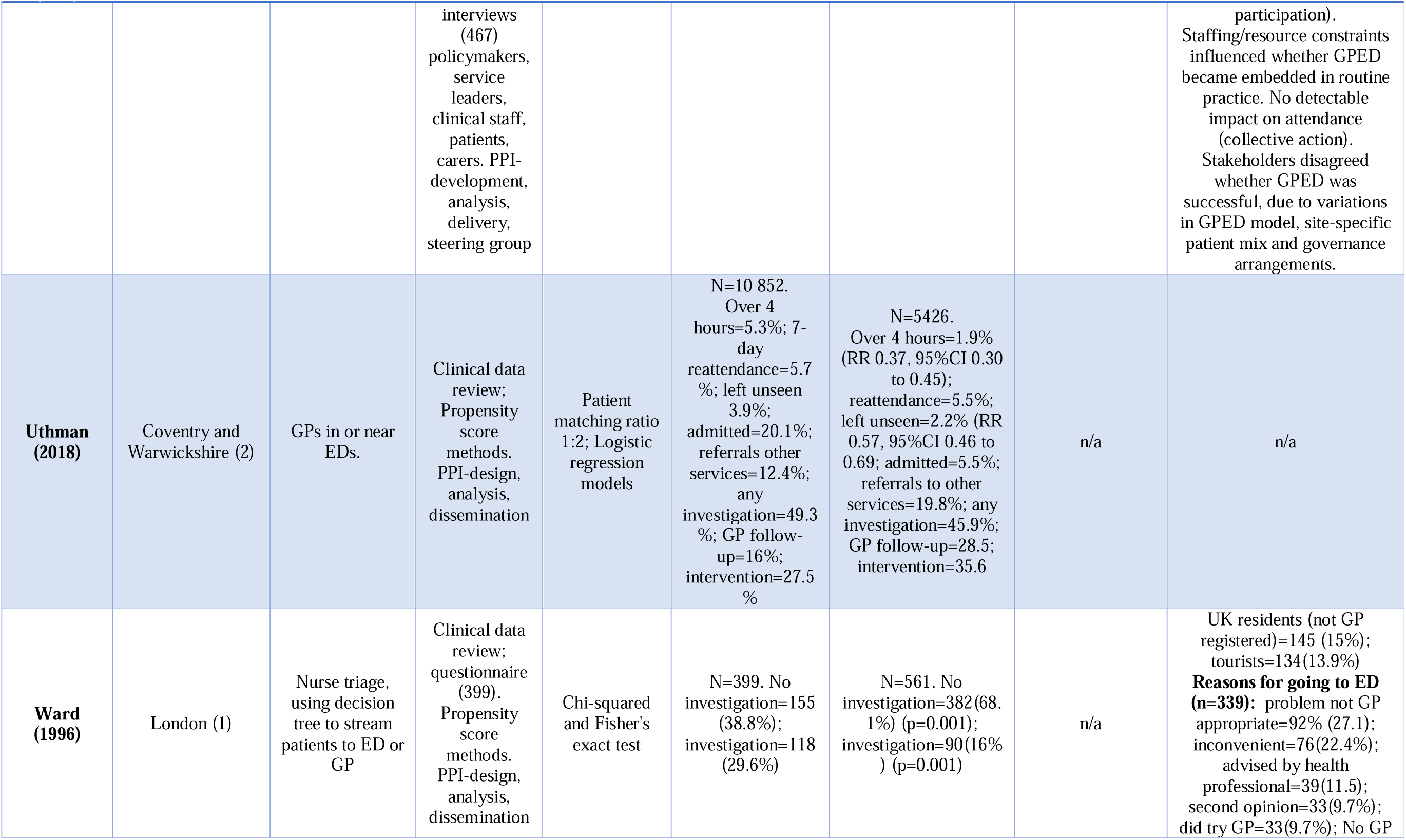
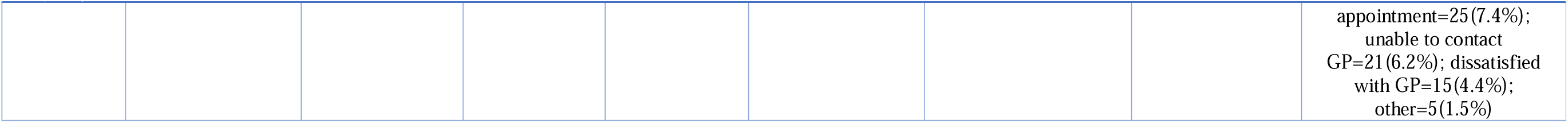

Seven publications (Anderson et al., 2021; Brant et al., 2021; Edwards et al., 2020; Edwards et al., 2022; Edwards et al., 2021; Gaughan et al., 2022; Scantlebury et al., 2022) reported different perspectives of primary care services within EDs that originated from a large, multi-strand project across ten sites in England (Benger et al., 2022). Two studies used the same data for different analyses (Chalder et al., 2007; Salisbury et al., 2007).

### Patient and public involvement

Five of the seven studies originated from a large, multi-strand project across ten sites in England (Benger et al., 2022), and reported PPI (Brant et al., 2021; Anderson et al., 2021; Edwards et al., 2020; Edwards et al., 2021; Edwards et al., 2022). Ten people formed the stakeholder PPI group for the project and contributed to all parts of the research (design, tool development, analysis, theory development, dissemination). Uthman et al. (2018) and Ward et al. (1996) also included PPI in research design, analysis, and dissemination.

### Models used for service delivery

Some taxonomies have been applied to describe the care delivery models available. Ablard et al. (2017) described three models in 13 hospitals in Yorkshire and Humber : primary care embedded in ED (n=7); Co-located centres (n=2); GP out-of-hours located next to EDs (n=9, of which 5 sites concurrently offered primary care-embedded in ED). Brant et al. (2021) described how delivery formats changed between 2017 and 2019. They proposed a 4-group taxonomy: 1) *inside/integrated,* 2) *inside/parallel,* 3) *outside/onsite,* 4) *outside/offsite*. Brant et al. (2021) observed that inside/parallel primary care services increased by 49% in 2017- 2019, while ‘Inside/integrated’ models fell to just 9% of initiatives that included GP services in or alongside EDs in England. They suggest these changes transpired after funding became available to separate primary care services from EDs. Neither Ablard et al. (2017) nor Brant et al. (2021) presented data to assess effectiveness of these diverse models.

### Types of streaming

Edwards et al. (2021) developed a taxonomy to describe four streaming systems of patients in ten hospitals based on where streaming took place and who did it. Their divisions were: 1) *Front door streaming* (patients streamed by a nurse at the front of the emergency department – before being booked in at reception), 2) *Streaming inside the emergency department* (patients streamed by a nurse working inside the emergency department– after being booked in at ED reception), 3) *No primary care streaming* (a traditional style triage, with GPs using clinical judgement to select patients they would treat) 4) *Combined streaming pathways* (combinations of 1–3 within the emergency department or across the ED and primary care services, varying at different times). Streaming could and sometimes was mixed with triage, leading to staff disquiet about what their clinical objectives should be at specific points in the care pathway (Anderson et al., 2021). Other studies echoed these findings about the inconsistent service objectives at specific pathway points (Ablard et al., 2017; Aldus et al., 2022; Edwards et al., 2020; Edwards et al., 2022; Jones, 2011; Scantlebury et al., 2022).

### Clinical outcomes

Ten studies provided reportable data, replicated in Table 5.

**Table 5:** Primary care and emergency department clinical outcomes of included studies for the systematic review on primary care clinicians working in or near Emergency Departments.

| Study | Wait time under 4 hours-time (%) |  | Investigations % |  | Referrals<sup>1</sup> % |  | Prescriptions % |  | Left without being seen % |  | Unplanned reattendance % |  |
| --- | --- | --- | --- | --- | --- | --- | --- | --- | --- | --- | --- | --- |
|  | PC | ED | PC | ED | PC | ED | PC | ED | PC | ED | PC | ED |
| **Aldus 2022** | 1.5 hours (100) | - | 26.6 | - | 64.2 | - | - | - | - | - | - | - |
| **Dale 1995** | - | - | 17 | 38.3 | 93.2 | 84 | 37.6 | 39.5 | - | - | - | - |
| **Gaughan 2022** | No impact | (83.5) | - | - | - | - | - | - | No impact | 2.36 | No impact | 9.1 |
| **James 2019** | 1.25 hours | 2.75 hours | 1.48 | 5.57 | - | - | 17.6 | 36.8 | - | - | - | - |
| **Jones 2011** | 3 hours (100) | - | - | - | 25* | - | - | - | 2* | - | - | - |
| **Leigh 2021** | 0.66 hours (88.4) | 2.75 hours (88.4) | - | - | 30.3 | 17.6 | 15.1 | 10.8 | 1.2 | 9.7 | - | - |
| **McCarron 2019** | 1 hour | 1.8 hours | 7.2 | 7.6 | - | - | 34.3 | 34.9 | - | - | 7.1 | 9.7 |
| **Salisbury 2007** | 1.8 hours (95.6) | 2.4 hours (94.6) | - | - | - | - | - | - | - | - | - | - |
| **Uthman 2018** | (98.1) | (94.7) | 45.9 | 49.3 | 32.5 | 25.3 | - | - | 2.2 | 3.9 | 5.5 | 5.7 |
| **Ward 1996** | - | - | 16 | 29.6 | - | - | - | - | - | - | - | - |

*Note:* <sup>1</sup> Includes referral to other services and admission rates; PC-primary care; ED-emergency department; \* approximate values, please appendix 4 for detailed values; - no value reported in the study

### Wait time

Eight studies assessed wait times. Most patients were seen or treated within four hours, irrespective of the site they visited. However, patients in ED streams waited longer than those in primary care streams. Leigh et al. (2021) reported that patients waited less than an hour in primary care, but 2.75 hours in ED. Similarly James et al. (2019) reported 1.25 hours wait in primary care and 2.75 in ED.

### Investigations conducted

Six studies provided details of investigations. One study presented primary care stream data only (Aldus et al., 2022). Of the five studies providing comparisons between primary care and EDs, only two presented significant differences in patients having investigations. Dale et al. (1995) showed 38.3% of patients went for investigations by ED compared to 17% in primary care. Similarly Ward et al. (1996) reported 29.6% (ED) vs 16% (primary care).

### Referral rates

Five studies provided data on referral of patients to other services or hospital admittance. Aldus et al. (2022) only presented referral information for primary care services (64.2%). An approximate referral rate of 25% for primary care services was extrapolated for Jones (2011). All comparison studies showed greater referral rates for primary care compared to ED pathways, Dale et al. (1995) reported 93.2% vs 84%, Uthman et al. (2018) reported 32.5% vs 25.3% and Leigh et al. (2021) reported 30.3% vs 17.6%.

### Prescription rates

Of the four studies providing prescription rates, only James et al. (2019) presented significant differences in prescription rates between primary care (17.6%) and ED (36.8%) pathways.

### Left before being seen

Two studies compared how many patients left primary care or ED pathways before being seen data. Leigh et al. (2021) reported 9.7% of people left ED compared to 1.2% leaving primary care before being seen, while Uthman et al. (2018) reported similarity between the pathways (2.2% versus 3.9%).Two studies compared how many patients left primary care or ED pathways before being seen data. Leigh et al. (2021) reported 9.7% of people left ED compared to 1.2% leaving primary care before being seen, while Uthman (2018) reported similarity between the pathways (2.2% versus 3.9%).

### Other outcomes

Salisbury et al. (2007) found that patients were more likely to be seen by a nurse in walk-in centres (WICs) but also concluded that their evaluation of WICs was hampered because the WIC model was limited in the hospitals included in the study, thus limiting comparison of attendance rates, process, outcomes, and costs. Similar findings were in Coleman et al. (2001). Gaughan et al. (2022) analysed a dataset of 4.4 million attendances to 40 English EDs from April 2018 to March 2019, concluding that GP services in these EDs had had no or negligible impact on these outcomes: percentage of patients discharged within 4 hours of arrival; ED attendances that resulted in hospital admission; patients who left without being seen; unplanned reattendance at the ED within 7 days; 30-day mortality; non-urgent ED attendances; volume of ED attendances.

### Adverse clinical outcomes

No adverse outcomes were reported. Scantlebury et al. (2022) suggested streaming staff may err on the side of caution when patients are streamed to primary care, while Aldus et al. (2022) reported that staff felt that they were being required to be too cautious about who could be streamed to GP care.

### Patient experience

There was little change in the number of patients attending walk-in centres (WIC) rather than EDs, even when they were located nearby. Chalder et al. (2007) found that patients (79%) with injuries attended ED first and 55% were unaware they had been redirected to WIC. Most patients found both pathways convenient. Most patients were satisfied with the care they received, but more people were dissatisfied with the ED due to duration of visit, facility cleanliness, time to discuss healthcare, involvement in decision making and privacy (Chalder et al., 2007).

Similarly, Coleman et al. (2001) found that many patients (62%) were unaware of other services and considered availability of other services problematic (38%). Patients (48%) thought they needed a radiograph and other people (43%) advised patients to attend ED. Overall patients (70%) had a positive experience in ED and 76% perceived their health condition as serious, while 38% wanted reassurance.

Ward et al. (1996) reported that 15% of UK residents attending an ED in London were not registered with a GP, and almost 14% more attendees were tourists. Reasons patients gave for attending ED included the health condition was considered serious and required ED attention (27.1%), convenient to go to ED (22.4%), they were advised by healthcare professional (11.5%), problems with getting GP appointments (13.6%), and/or had dissatisfaction with GP (4.4%).

Aldus et al. (2022) found patients were satisfied with the wait time and treatment they received in primary care situated in or near an ED, however, they had no comparator data from patients treated in the co-located ED. Dale et al. (1996) found high levels of patient satisfaction for all aspects of primary care or ED consultation: satisfied with clinical assessment (77%); happy with treatment (75%); happy with consulting doctor’s manner (88%).

### Staff perceptions

Overall, there were diverse views about the utility of primary care in or near EDs, possibly due to variations in the models, patient variations, and governance provision (Scantlebury et al., 2022). Some staff queried the justification for using primary care within EDs (Ablard et al., 2017; Scantlebury et al., 2022). Perceived challenges and facilitators of primary care in or near EDs, included staff engagement, governance, space, recruitment, training, funding, and service demand (Edwards et al., 2020; Scantlebury et al., 2022). It was felt that primary care in ED improved patient flow and allowed clinicians to spend more time with acute patients, but it also posed the risk of encouraging patients to attend ED (Aldus et al., 2022; Jones, 2011). The wellbeing of staff in Norwich was unaffected by the new way of working and they felt satisfied due to the streamlined service and reduced patient waiting, however, they were frustrated by the working environment and refusal of some departments to co-operate with GP staff (Aldus et al., 2022).

### Economic outcomes

Three studies presented costs of primary care versus ED care (Leigh et al., 2021; Dale et al., 1996; Salisbury et al., 2007). Each study evaluated costs differently, however, only one concluded that primary care cost less than ED, due to differences between clinician salaries (Dale et al., 1996), Table 6

**Table 6:** Economic evaluations presented in studies included in the systematic review about primary care in or near Emergency Departments.

| Study | Cost of primary care per patient | Cost of ED care per patient |
| --- | --- | --- |
| Dale et al. 1996 | £11.70 | £18.64 |
| Leigh et al. 2021 | £115.24 | £141.16 |
| Salisbury et al. 2007 | £117.18 | £121.67 |

### Quality assessment

The summary of Quality assessment is in Table 7. Results from applying the MMAT (MMAT; Hong et al., 2018) showed an initial 75% agreement between reviewers. After consensus was reached quality assessments scores were applied to define high quality studies to be those with four or five ‘yes’ responses and lower quality studies were those ≤ 3 ‘yes’ responses. All five qualitative studies and three out of five mixed methods studies (Aldus et al., 2022; Coleman et al., 2001; Scantlebury et al., 2022) were deemed to be high quality (5/5) (refs), while Brant et al. (2021) and Jones (2011) were low quality (3/5). Five of the quantitative studies were considered high quality (4/5) (Uthman et al., 2018; Leigh et al., 2021; James et al., 2019; Ward et al., 1996; Gaughan et al., 2022), and five were low quality. Dale et al. 1995/1996 and McCarron et al. (2019) scored three and Salisbury et al. (2007) scored just two. Overall, 13 studies were deemed to be good quality.

**Table 7:**
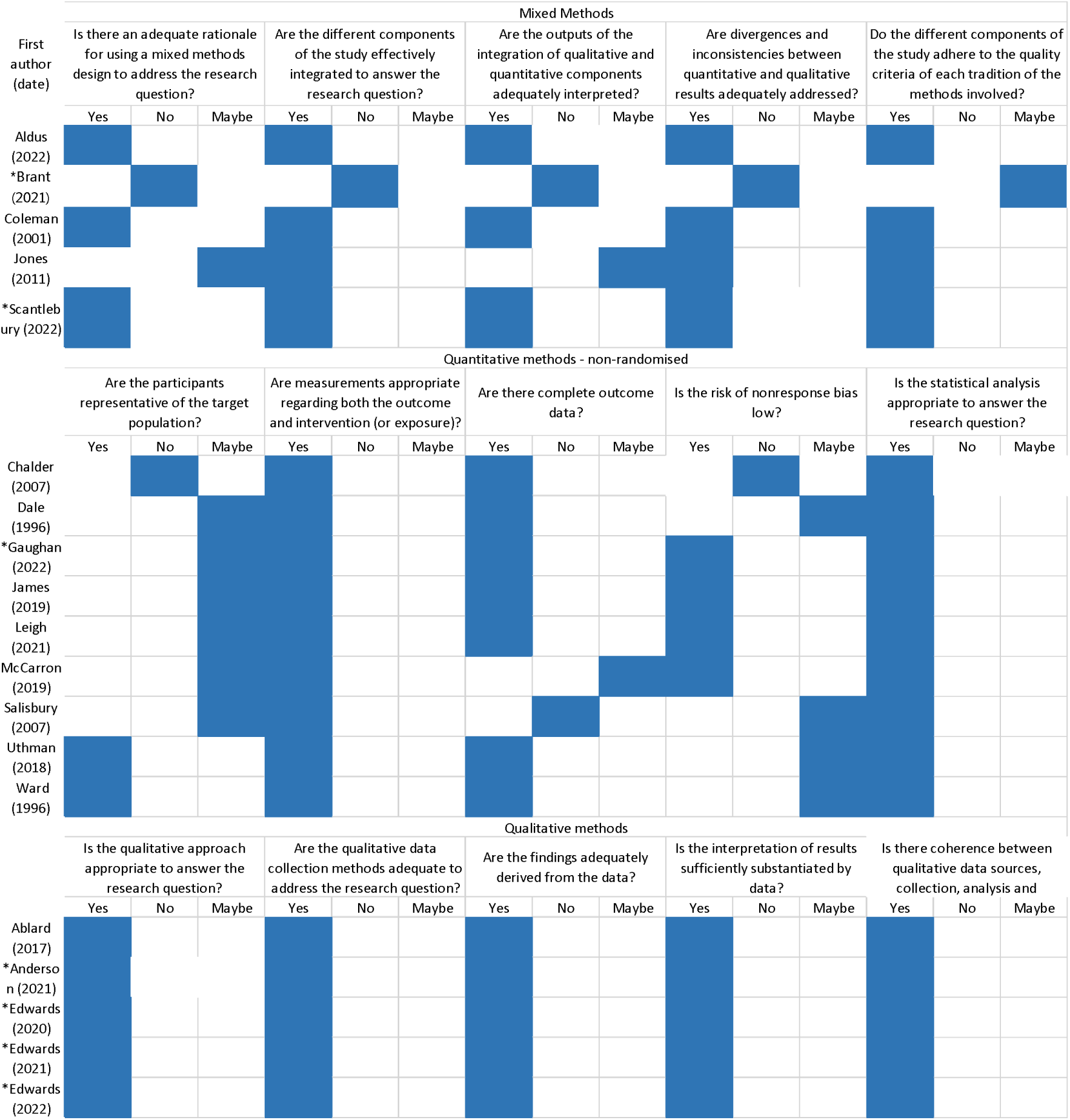
Mixed Method Appraisal Tool quality assessment of included studies for the system<u>atic review on primary care in or near Emergency Departments</u>.

## Discussion

Twenty diverse studies assessed various aspects of primary care services in or near EDs. Our review provided a detailed focus of research conducted thus far on the topic and indicates that primary care services in or near EDs have little clinical effect on target outcomes and are not cost effective.

Conclusions in the international scoping review by Jeyaraman (2021) were positive about potential benefits of deploying primary care professionals to reduce ED overcrowding. The difference between their positive conclusions and our equivocal findings is partly because we confined our evidence summary to when primary care professionals specifically provided treatment, and only included studies conducted in the UK. Our divergent findings may also arise from the heterogeneity of the research and the small number of (UK) studies available at present. Our conclusions concur with seven studies arising from a single UK-focused project, in which no clear net benefits for providing primary care at EDs were found (Benger et al., 2022). Many deficiencies can be easily identified in the existing literature. Wait times and indeed many other outcomes are only reported relatively. Reporting on adverse outcomes was not thorough and data about patient outcomes after attendance were missing. There is a lack of obvious research about whether such initiatives especially help vulnerable communities and as such reduce inequalities in access to health care. Whether they have the potential to contribute to widen inequalities warrants further research.

How primary care is incorporated into EDs raised concerns from staff. Many staff expressed concern over whether there is an actual need for primary care services in EDs, criticised how the service is integrated, how patients are streamed and whether they are effective and cost-effective (Ablard et al., 2017; Aldus et al., 2022; Anderson et al., 2022; Brant et al., 2021; Edwards et al., 2020; Edwards et al., 2022; Edwards et al., 2021; Scantlebury et al., 2022).

There were also practical issues around the working environment, infrastructure, governance, communication, poor working relationships and staff recruitment (Aldus et al., 2022; Anderson et al., 2022; Edwards et al., 2020; Edwards et al., 2022; Edwards et al., 2021; Jones, 2011). Yet, some staff acknowledged the benefits of having primary care staff working alongside ED clinicians because there is a sharing of information, different way of diagnoses and learning opportunities (Ablard et al., 2017; James et al., 2019). There is an under- exploited opportunity when primary care clinicians are working in parallel to emergency department staff to compare diagnostic and risk management strategies deployed by the respective types of clinicians and which one is more appropriate for low acuity presentation patients.

Research should assess how best to implement primary care services in or near EDs for effective delivery of clinical outcomes. Also, which primary care clinicians are best to deliver the care. Nurses already have a role in WICs (Salisbury et al., 2007) and research could assess different diagnostic and treatment regimens of each pathway. The potential scope for harms or benefits are much wider than individual patient experiences, though. For instance, there is potential harm if diagnosis and treatment are delayed for high-risk patients, and potential benefit if other conditions are treated quickly. Also, offering primary care at acute hospital settings may undermine demand for and quality of care received at conventional GP services (Anderson et al., 2022). Research should also provide comprehensive cost-benefit analyses to help clarify the benefits and risks of primary care services in or near EDs.

A diversity of reasons was described for why patients attended ED rather than take the NHS preferred route through primary care (Aldus et al., 2022; Chalder et al., 2007; Coleman et al., 2001; Dale et al., 1995; Ward et al., 1996). The 4-hour wait at ED was acceptable and few rated difficulties in getting a GP appointment as a reason to go to ED. Shorter wait times for patients in primary care was advocated by the patient advisors in our research and this may be more generally appreciated. Patients were satisfied with the service they received, irrespective of which service pathway. Better understanding of why patients attend ED rather than attend primary care services might support development of behaviour change programmes to encourage patients to persist on more appropriate pathways, including attendance to their registered GP surgery but also care-seeking at pharmacies, telephone consultations and walk-in centres.

Stratification of outcomes by patient type (gender, age, socio-economic status) was not possible in the included studies; in reality, patient health care seeking behaviours may well vary with these demographic traits. Process evaluation (Meysman et al., 2022) may be required to truly understand which aspects of GP in or near ED services have the best chance of providing the most benefits, to both patients and health care systems. Over time, an optimal service delivery format may emerge. It may be that prior GP-in or near-ED implementations should be seen as part of a natural evolution and selection process to arrive at an optimal service delivery format.

## Limitations

Our review only included studies looking specifically at primary care in or near EDs and included only UK studies. These criteria were adopted for the purpose of providing a holistic assessment of these NHS services but could also result in exclusion of studies that are somewhat similar and could offer additional information on primary care in or near EDs.

Despite a comprehensive search strategy, it is possible that some studies were not located or available.

## Conclusion

Overall, the evidence collected in this systematic review suggests that primary care clinicians providing treatment in or near EDs has had negligible effect (good or bad) on clinical outcomes, or economic evaluations. Clinicians have mixed opinions about the effectiveness of siting primary care services in or near EDs. Patients are generally satisfied with the healthcare they receive and do not seem to be aware of the different care pathways. This may be due to the general lack of consensus in service provision about what constitutes primary care in or near EDs.

## Author contributions

JB and SH conceived the systematic review. JB and SHW ran the searches, screened articles, extracted and analysed the data. SH, PE and DP provided expert guidance on qualitative papers and clinical roles and refereed any disagreements. SHW and JB wrote the first draft and assembled revisions. All authors revised for content.

## Data Availability

All data produced in the present study are available upon reasonable request to the authors

